# Effectiveness of one dose of MVA-BN vaccine against mpox infection in males in Ontario, Canada: A target trial emulation

**DOI:** 10.1101/2023.10.04.23296566

**Authors:** Christine Navarro, Cindy Lau, Sarah A. Buchan, Ann N. Burchell, Sharifa Nasreen, Lindsay Friedman, Evaezi Okpokoro, Peter C. Austin, Darrell HS Tan, Jonathan B. Gubbay, Jeffrey C. Kwong, Sharmistha Mishra, Canadian Immunization Research Network (CIRN) Provincial Collaborative Network Investigators

## Abstract

**Background:** Limited evidence is available on the real-world effectiveness of modified vaccinia Ankara-Bavarian Nordic vaccine (MVA-BN) against mpox infection.

**Methods:** We emulated a target trial using linked databases in Ontario, Canada to estimate the effectiveness of one dose of MVA-BN. Our study included males aged ≥18 years who: (1) had a history of syphilis testing and a laboratory-confirmed bacterial sexually transmitted infection (STI) in the prior year; or (2) filled a prescription for HIV pre-exposure prophylaxis in the prior year. On each day between June 12, 2022 and October 27, 2022, males who had been vaccinated 15 days prior were matched 1:1 with unvaccinated males by age, geographic region, prior HIV diagnosis, number of bacterial STI diagnoses in the previous three years, and receipt of any non-MVA-BN vaccine in the previous year. We used a Cox proportional hazards model to estimate the hazard ratio comparing the incidence of mpox between the two groups, and calculated vaccine effectiveness as (1–HR)x100.

**Results:** Each group included 3,204 males. A total of 71 mpox infections were diagnosed over the study period, with 0.09 (95% confidence interval [CI], 0.05–0.13) per 1000 person-days for the vaccinated group and 0.20 (95%CI, 0.15–0.27) per 1000 person-days for the unvaccinated group. Estimated vaccine effectiveness of one dose of MVA-BN against mpox infection was 59% (95%CI, 31–76%).

**Conclusions:** This study, conducted in the context of a targeted vaccination program and evolving outbreak, suggests that one dose of MVA-BN is moderately effective in preventing mpox infection.

## INTRODUCTION

In May 2022, more than 20 countries where mpox had not been previously identified reported cases to the World Health Organization.^1^ On July 23, 2022, the global mpox outbreak was declared a public health emergency of international concern, and targeted use of second- or third- generation smallpox vaccines for outbreak control was recommended.^2^

Modified vaccinia Ankara vaccine (MVA-BN [Bavarian-Nordic]; trade names Imvamune®, Jynneos®, Imvanex®) is a third-generation, live-attenuated, non-replicating smallpox vaccine.^3^ In Ontario, Canada, MVA-BN was deployed in June 2022 as post-exposure prophylaxis for high-risk contacts (but few doses were given in this context) and pre-exposure prophylaxis for gay, bisexual, and other men who have sex with men (gbMSM), and sex workers at high risk of exposure to mpox.^4^ Although MVA-BN is approved in Canada as a series of two doses 28 days apart, Ontario initially employed a dose-sparing strategy such that vaccine candidates could only receive one dose due to concerns about limited vaccine supply. A two- dose (0.5 mL, subcutaneous) program was subsequently implemented on September 30, 2022.

Prior to the global mpox outbreak, limited clinical or real-world data on the use of MVA- BN to prevent mpox infection existed.^5,6^ Estimates of the effectiveness of a single dose of MVA- BN obtained using various observational study designs have since emerged in the literature, ranging from 36% to 86%.^7–13^ As with all observational studies, each report has discussed the potential for residual confounding and selection biases. Only one study to date emulated a target trial in an effort to address these biases, but it was restricted to HIV-negative males taking HIV pre-exposure prophylaxis.^12^ The objective of the current study was to estimate vaccine effectiveness of one dose of MVA-BN against laboratory-confirmed mpox infection through a target trial emulation.

## METHODS

### Study design, setting, and population

We conducted a target trial emulation to answer the causal question of interest (**Figure S1**) and minimize biases, particularly confounding.^14^ We used laboratory, vaccination, reportable diseases, and health administrative data from Ontario (population 15.1 million as of July 2022), which has a single-payer healthcare system. All datasets included in the analysis (detailed in the **Supplementary Methods**) were linked using unique encoded identifiers and analyzed at ICES (formerly the Institute for Clinical Evaluative Sciences). Ethical approval for this study was obtained from Public Health Ontario’s Ethics Review Board.

The study period captured the beginning of pre-exposure vaccine availability (June 12, 2022 to November 26, 2022; **Figure 1**). The end date was chosen based on percent positivity being 0% for two consecutive weeks and the last outbreak-associated mpox case being reported on November 10, 2022.^15^ Eligibility for single-dose, pre-exposure vaccination comprised gbMSM reporting one or more of the following: diagnosis of bacterial sexually transmitted infection (STI) in the previous two months; currently engaging in or anticipating sex with two or more sexual partners; attending sex-on-premises venues; or engaging in anonymous sex. Pre-exposure vaccination eligibility also included individuals engaged in sex work, immunocompromised individuals, or pregnant individuals if they were contacts of persons at risk, as defined above.^16^

**Figure 1.**
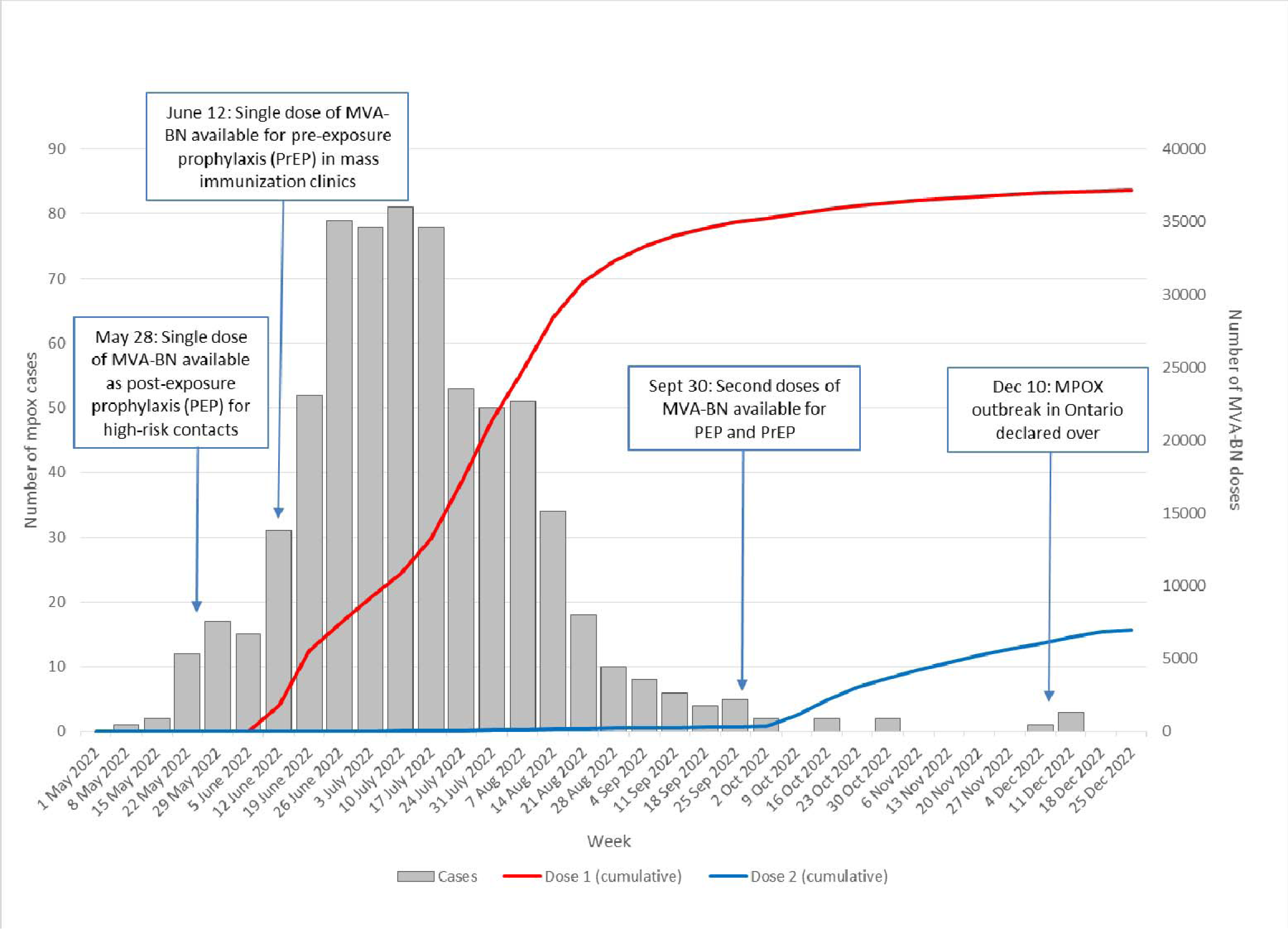
Confirmed mpox cases and number of modified vaccinia Ankara-Bavarian Nordic (MVA-BN) vaccine doses administered by week: Ontario, Canada, May 1 to December 31, 2022.

Eligibility criteria for the target trial were conceptualized to minimize confounding stemming from the correlation between risk of infection and vaccination. The study population was restricted to males aged ≥18 years as of June 12, 2022 with at least one of the following proxies for risk of exposure to mpox as of the date of enrolment (between June 12, 2022 and October 27, 2022, to ensure each person could have at least 30 days of observation): (1) at least one syphilis test in the previous year and a new diagnosis of one or more bacterial STI (gonorrhea, chlamydia, or syphilis) in the previous year; or (2) a filled prescription for HIV pre-exposure prophylaxis (PrEP) in the previous year (all definitions provided in **Table S1**). We excluded individuals with a documented positive mpox polymerase chain reaction (PCR) test prior to June 12, 2022.

### Intervention and outcome

The intervention of interest was vaccination with a single dose of MVA-BN. We were unable to estimate the effectiveness of a second dose due to few individuals having received a second dose (13.7% of those who received one dose) by the end of the study period (November 26, 2022) and few mpox cases being diagnosed in October and November. The outcome of interest was PCR-confirmed mpox infection. Based on immunogenicity data, an individual was classified as vaccinated >14 days after the first dose.^17^

### Specification and emulation of the target trial

On each day between June 12, 2022 and October 27, 2022, males who had been vaccinated with a single dose of MVA-BN 15 days prior were matched in a 1:1 ratio to unvaccinated controls. We followed individuals until the earliest date of any of the following events: outcome, death, receipt of a first vaccine dose (for unvaccinated controls), receipt of a second dose, or end of the study period. Newly vaccinated males could enter the study even if they had previously served as a control.

To balance the distribution of measured baseline covariates that are associated with both the probability of vaccination and of mpox infection between vaccine recipients and controls, we matched vaccine recipients and controls with respect to: age (within 5 years), geographic region (since the epidemic trajectory and vaccine uptake varied regionally), proxies for sexual exposures (number of bacterial STIs in the previous three years, HIV status), and a proxy for vaccine confidence (history of receipt of any non-MVA-BN vaccine in the previous year). Details of the matching algorithm are provided in the **Supplementary Methods**.

We conducted three sensitivity analyses. To explore the potential for residual confounding by sexual risk exposures, we used two negative control outcomes that should not be directly affected by the receipt of MVA-BN but for which the effect of vaccination might be confounded.^18^ First, we measured the risk of mpox during the first 14 days after the first dose, when no difference between vaccinated and unvaccinated groups would be expected (“negative outcome period”). Second, we used a negative tracer outcome by estimating vaccine effectiveness against bacterial STI >14 days after vaccination; MVA-BN vaccine presumably has no benefit against acquisition of chlamydia, gonorrhea, or syphilis. However, STI diagnosis could be influenced by differential rates of testing following vaccination. Thus, we compared syphilis testing among vaccinated and unvaccinated groups over the study period to aid interpretation of the negative tracer outcome. Finally, we examined the potential for residual confounding related to socio-economic status by adjusting for neighborhood-level income, given that sexual networks and infection risks are shaped by systemic barriers to healthcare engagement and vaccine access.

### Statistical analysis

We examined covariate balance after matching using standard mean differences and considered a difference of ≥0.1 as potentially clinically meaningful. We estimated cumulative incidence functions (CIFs) for the vaccinated and unvaccinated groups and used a Cox proportional hazards model to estimate the hazard ratio (HR) comparing the hazard of mpox between the two groups. We calculated vaccine effectiveness as (1–HR)x100. Analyses were performed using SAS software, 9.4 (SAS Institute Inc., Cary, NC).

## RESULTS

A total of 9,803 males aged ≥18 years were eligible for the study. Of these, 3,204 who received the vaccine were matched to unvaccinated controls (**Figure 2**). The matched population was similar to the eligible population with respect to baseline characteristics (**Table S2**). All measured variables were well-balanced between the vaccinated and unvaccinated groups (**Table 1)**. The median age of matched participants was 35 years (interquartile range, 29-46) and more than half the participants were residents of Toronto.

**Figure 2.**
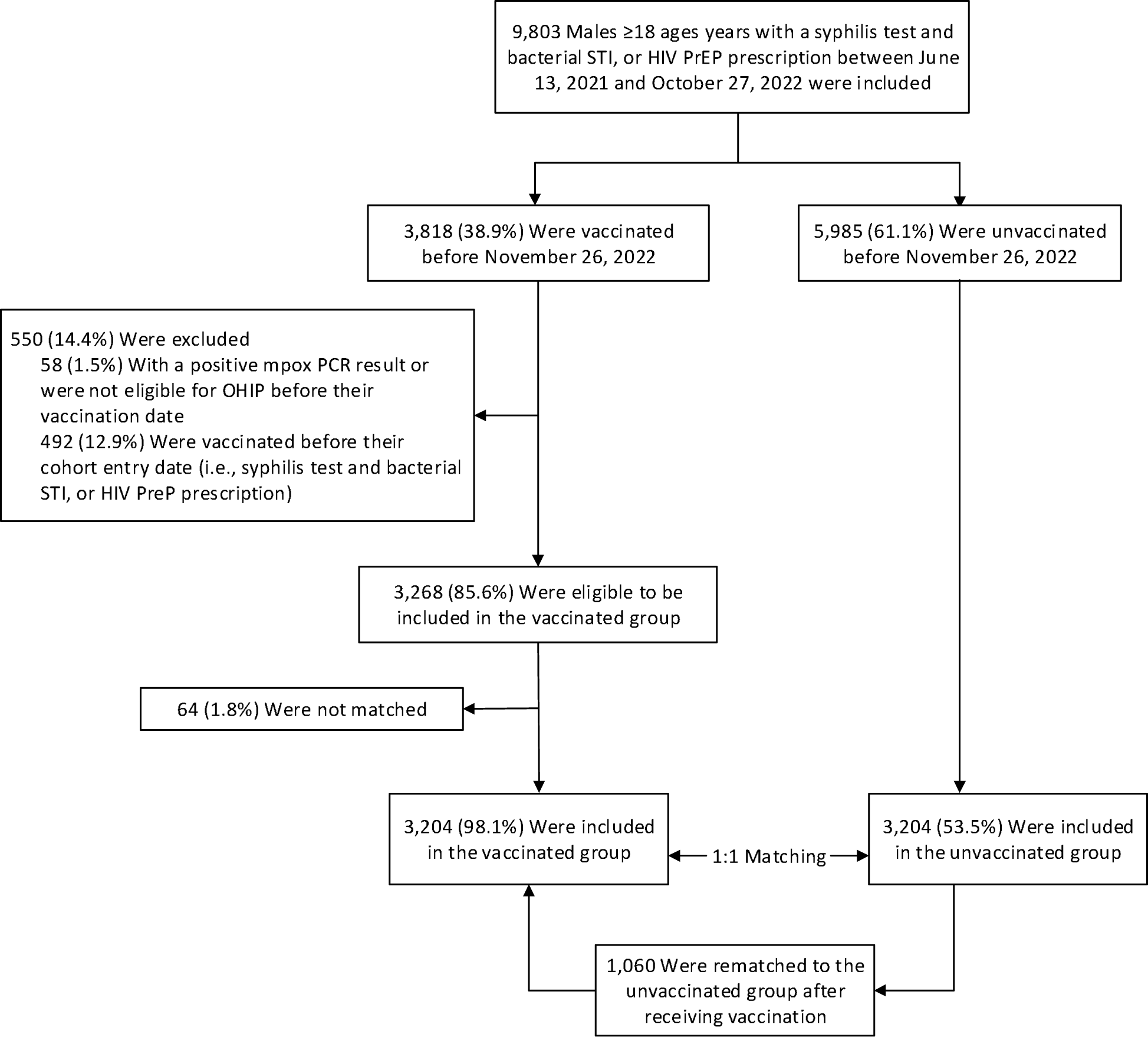
Cohort enrollment process and selection of persons for the emulation of a target trial evaluating effectiveness of the MVA-BN vaccine. A total of 9,803 males ages 18 years and older with a syphilis test and bacterial STI or HIV PrEP prescription between June 13, 2021 and October 27, 2022 (to allow at least 30 days of follow-up) were included in cohort. Of these, 6,408 persons (3,204 vaccinated persons matched to 3,204 unvaccinated persons based on birthdate within five years, geographic region, prior HIV diagnosis, number of bacterial STI diagnoses in the prior three years, and receipt of any other vaccine in the prior year) were included in the target trial. MVA-BN denotes Modified Vaccinia Ankara - Bavarian Nordic, STI sexually transmitted infection, HIV PrEP human immunodeficiency virus pre-exposure prophylaxis, PCR polymerase chain reaction, and OHIP Ontario Health Insurance Plan.

**Table 1.**
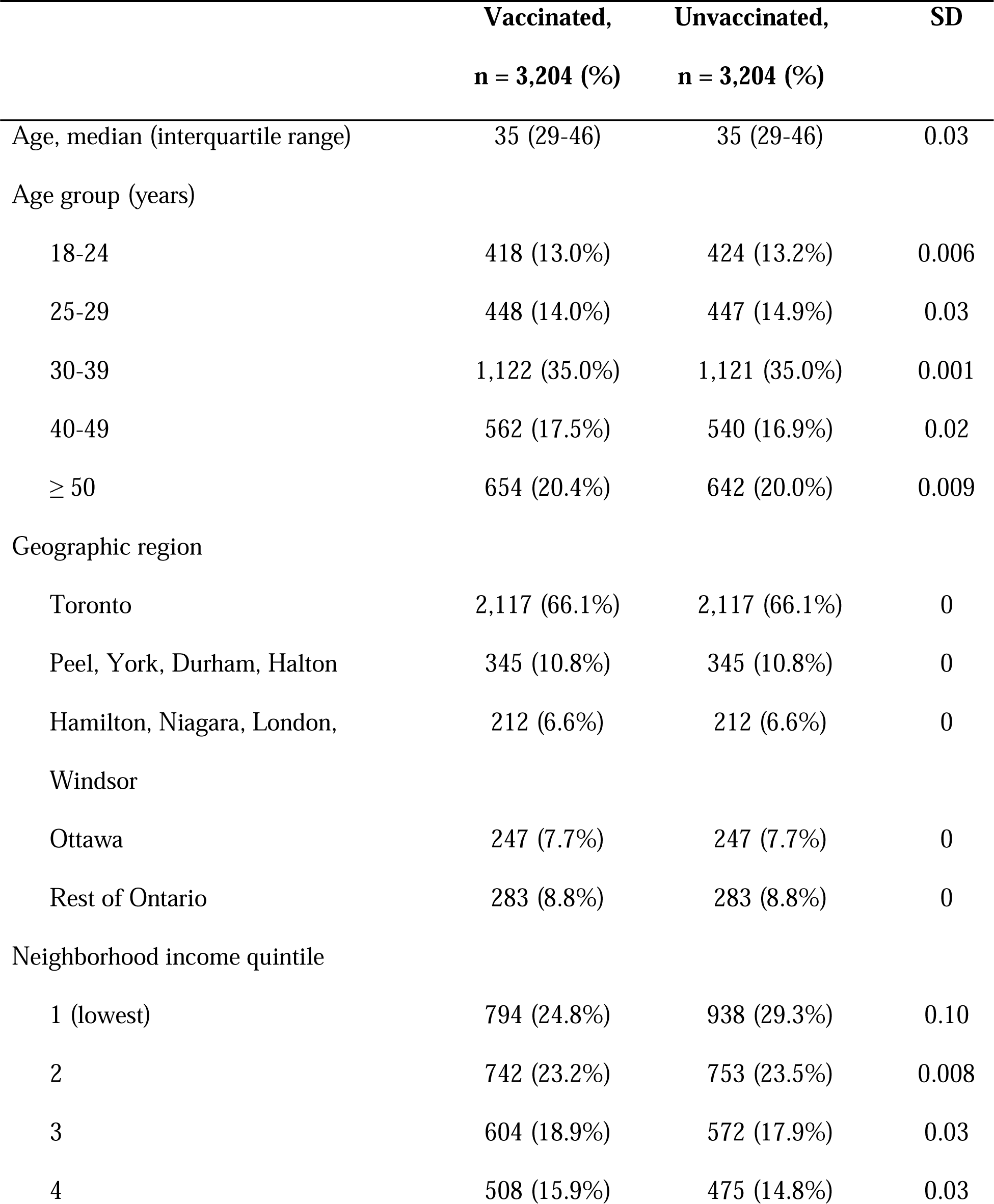
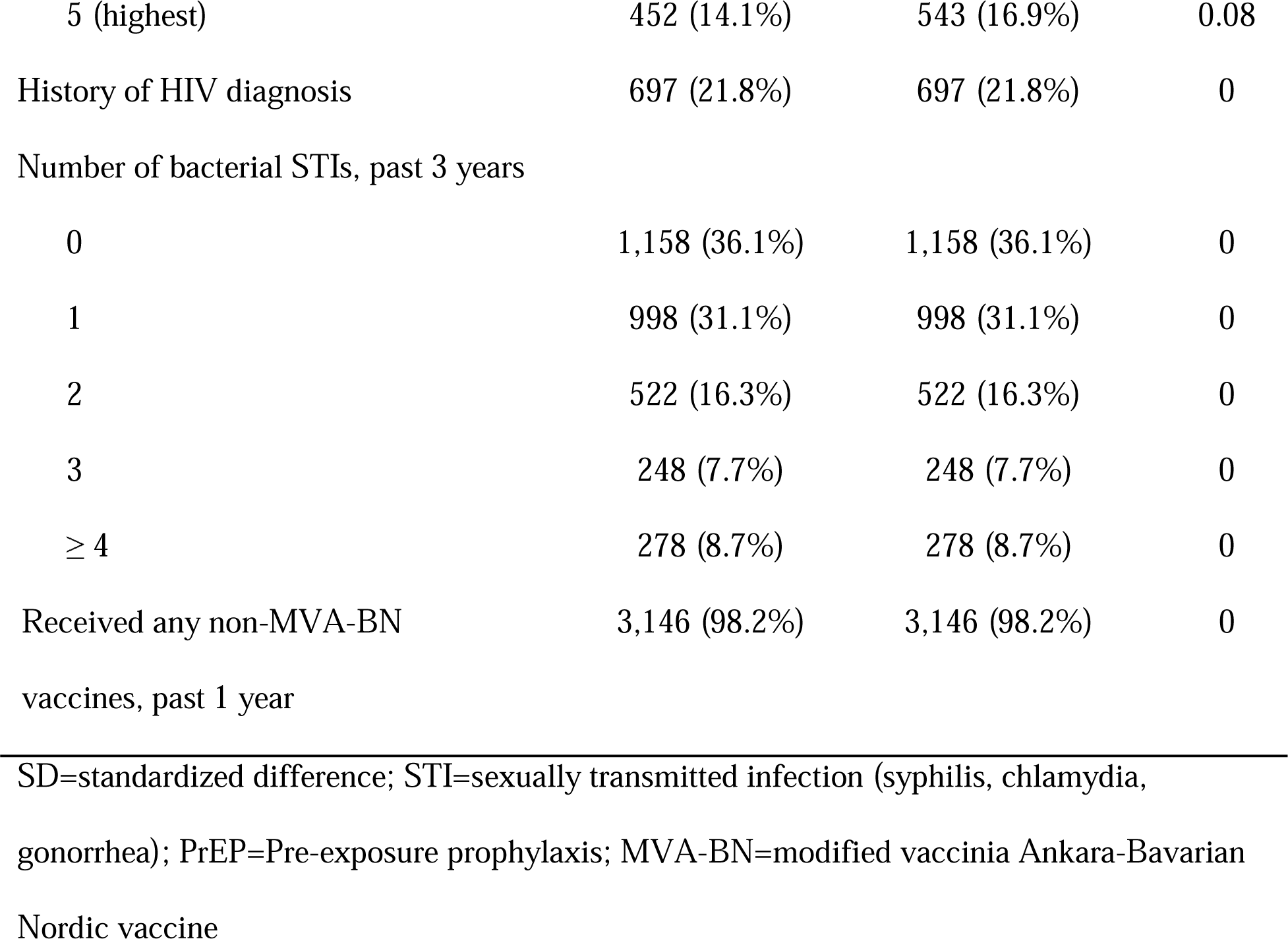
Demographic and sexual risk characteristics of the study population for the target.

During a median follow-up after the first dose of 85 days (interquartile range, 32–110) among vaccinated individuals, and 86 days (interquartile range, 31–111) among unvaccinated individuals, we observed a total of 71 infections, with 21 in the vaccinated group (0.09 per 1000 person-days; 95%CI, 0.05–0.13) and 50 in the unvaccinated group (0.20 per 1000 person-days; 95%CI, 0.15–0.27) over the study period of 153 days. We censored 293 (9.1%) individuals upon receipt of a second dose. **Figure 3** shows the cumulative incidence functions for the vaccinated and unvaccinated groups during the study period. The hazard ratio for vaccination was 0.41 (95%CI, 0.24–0.69), thus the estimated vaccine effectiveness for a single dose of MVA-BN against mpox infection was 59% (95%CI, 31–76%; **Figure 4**).

**Figure 3.**
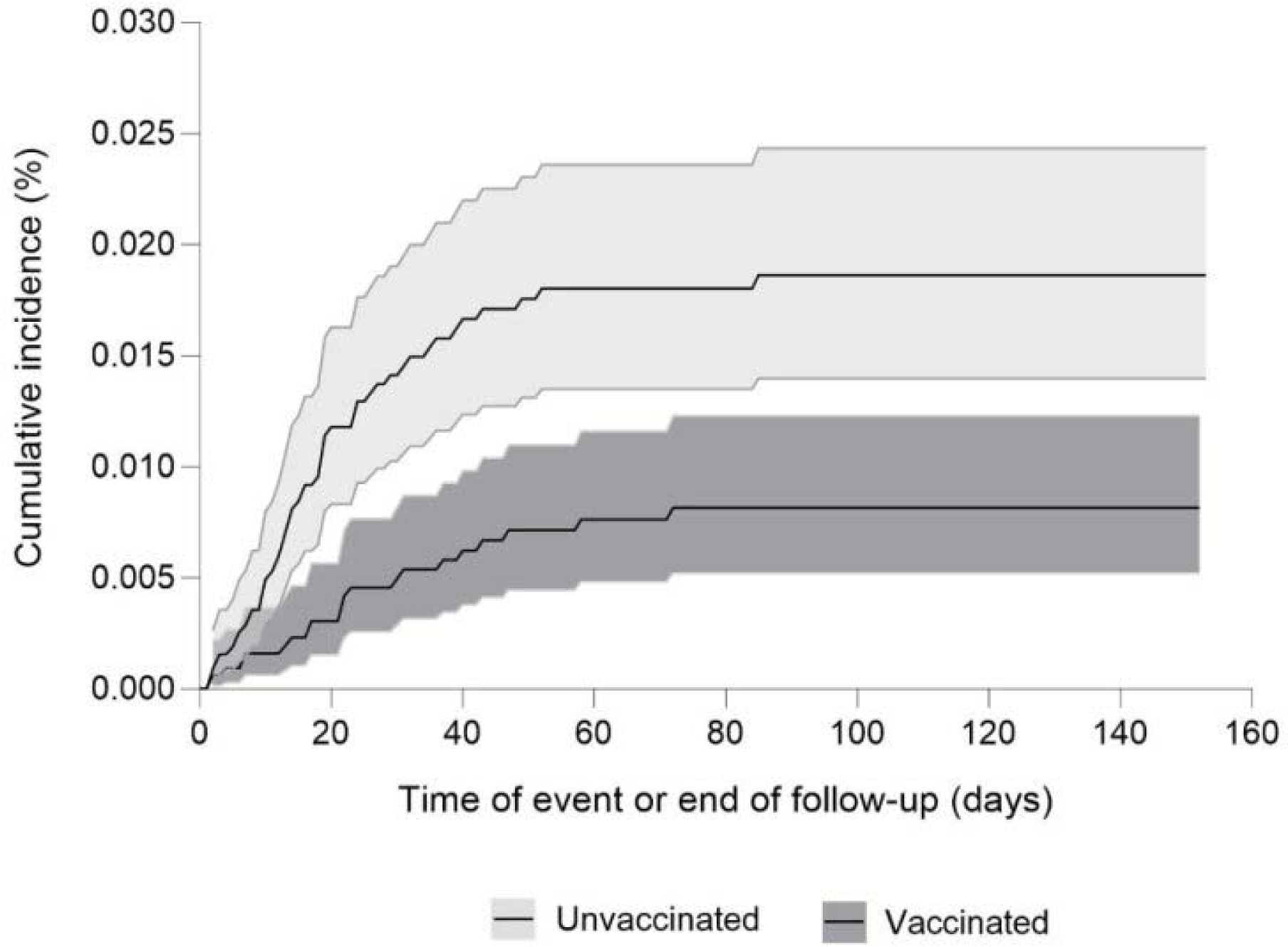
Cumulative incidence functions of confirmed mpox infection in Ontario, Canada, June 12, 2022 to November 26, 2022. Shaded areas represent 95% confidence intervals.

**Figure 4.**
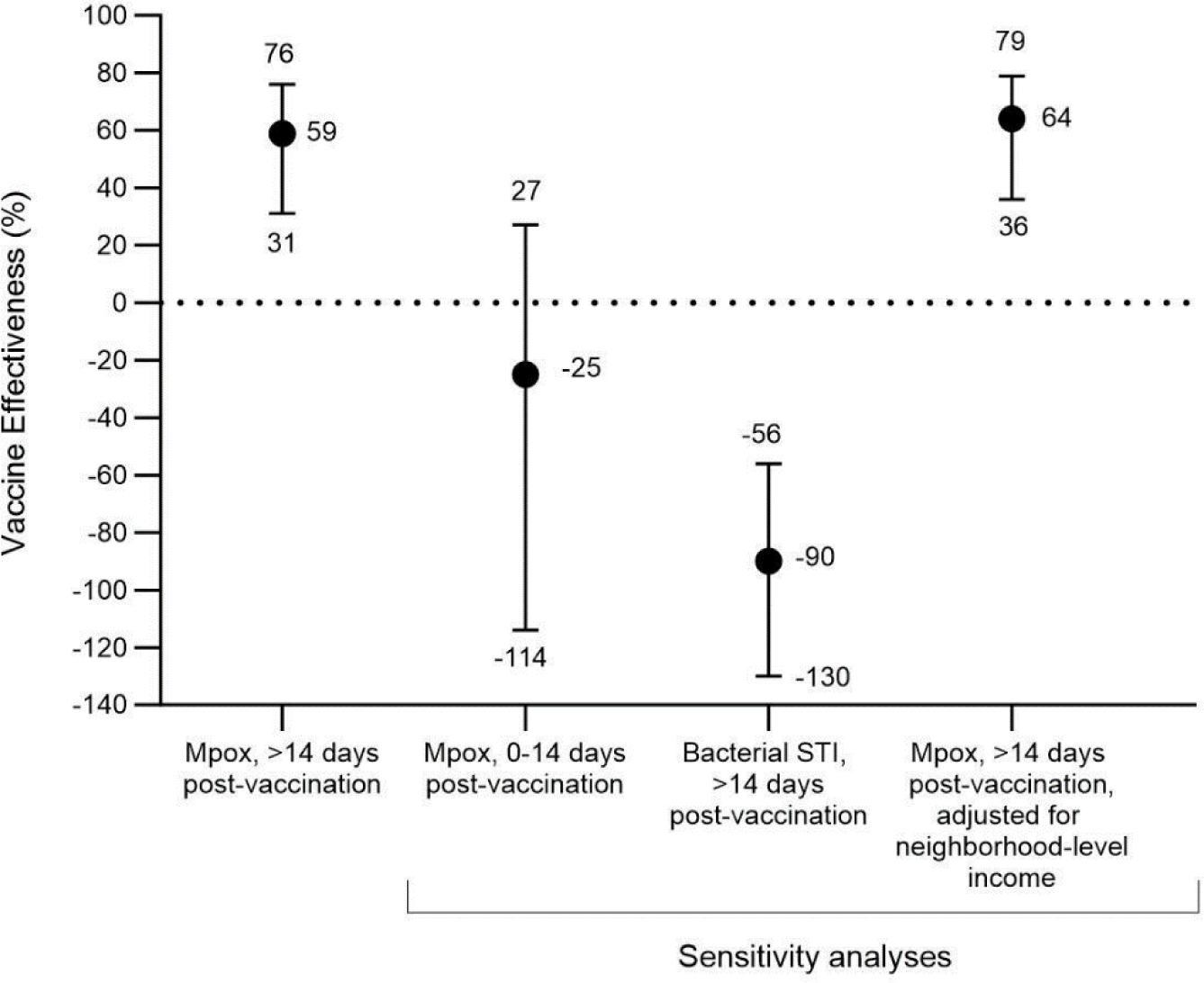
Estimates of vaccine effectiveness of one dose of MVA-BN between June 12, 2022 and November 26, 2022 in Ontario, Canada, primary and sensitivity analyses. MVA-BN denotes Modified Vaccinia Ankara - Bavarian Nordic, STI sexually transmitted infection.

Examination for residual confounding using negative outcomes demonstrated: (a) vaccine effectiveness of –25% (95%CI, –114%, 27%) during the first 14 days after receipt of vaccination; and (b) vaccine effectiveness of –90% (95%CI, –130%, –56%) against bacterial STI. Rates of first syphilis testing post-vaccination were 0.05 per 1000 person-days in the vaccinated group and 0.03 per 1000 person-days for the unvaccinated group. Finally, we did not identify a meaningful change in vaccine effectiveness against mpox after additionally adjusting for neighbourhood-level income (vaccine effectiveness 64%; 95%CI, 36–76%).

## DISCUSSION

Using a target trial emulation, we estimated the effectiveness of a single dose of MVA-BN vaccine to be moderate (59%; 95%CI, 31–76%) for preventing mpox infection in the context of a targeted vaccination program in Ontario, Canada. To confirm the specificity of this association, we determined that MVA-BN was not associated with a reduced rate of mpox infection during the first 14 days after vaccination (prior to developing an adequate antibody response) nor bacterial STI diagnoses (against which no protection would be expected).

Our estimate of vaccine effectiveness falls within the range observed across previous studies conducted in jurisdictions with similar epidemic dynamics and targeted vaccination programs. Our findings are most consistent with studies that restricted the study population to those at greatest risk of mpox exposure and that reduced time- and risk-based confounding.^7,9,10,12,13^ In the Canadian province of Québec, a test-negative study that used health administrative data and adjusted for exposure risks based on similar proxies as in our study (i.e., prior bacterial STI), estimated vaccine effectiveness against mpox infection to be 35% (95%CI, 2–59%).^13^ After further adjusting for self-reported measures of exposure risks (restricting analyses to those who completed a detailed questionnaire), vaccine effectiveness was estimated to be 65% (95%CI, 1–87%), similar to our estimate. Our estimate is lower than (but still compatible with) an estimate of 86% (95%CI, 59–95%) from a retrospective cohort study conducted in Israel that used more restrictive study eligibility criteria (i.e., living with HIV and a recent diagnosis of bacterial STI, or receipt of HIV PrEP),^9^ and an estimate of 79% (95%CI, 33–100%) from a target trial emulation conducted in Spain with even more restrictive study eligibility (i.e., enrolment restricted to males receiving HIV PrEP).^12^

Our study has several strengths. First, we used linked population-based databases within a publicly funded healthcare system to identify all MVA-BN vaccination events and all orthopox/mpox laboratory tests in Ontario. Second, to address the risks of residual confounding present in any observational study, we conducted rigorous matching across key potential confounders of the causal effect of vaccination on mpox infection. Risk-confounding is particularly important when estimating vaccine effectiveness because Ontario, like other jurisdictions, specifically targeted vaccination to individuals at greatest risk of infection. Evidence of exchangeability includes the similarity between groups with respect to proxies of sexual exposure risks, and similar outcomes during the “negative control period” before the vaccine was expected to confer protection. The negative tracer outcome analysis involving bacterial STI suggests that the observed vaccine effect is unlikely to be explained by differential reductions in sexual activity among gbMSM over the study period. In contrast, our finding of higher rates of newly diagnosed bacterial STI among vaccinated males suggests that the vaccination program successfully reached those most at risk of mpox and/or vaccinated males engaged in increased sexual activity post-vaccination. However, higher syphilis testing rates during the period following vaccination and higher STI rates among vaccinated individuals suggest additional STI testing opportunities and detection following engagement in preventive care with vaccination. Thus, residual risk-confounding resulting in an underestimation of vaccine effectiveness may be minimal. The study period included a rapidly evolving outbreak with mpox exposure risks declining quickly before a large fraction of study eligible population was vaccinated, thus there was a substantial risk for time-varying confounding due to differential exposure risks, which we mitigated by emulating a target trial.

Our study also has limitations. First, the rigorous matching meant that our final cohort comprised only 65% of the eligible population, with 71 outcomes for analysis. Although characteristics of included and eligible participants were similar, minimizing confounding came at a price of reduced sample size and precision. It also meant that subgroup analyses, such as among individuals aged >50 years, who may have received earlier generation smallpox vaccines, were not possible. Second, we were limited to routinely collected data, and information on history of previous smallpox vaccination, sexual exposures, and individual-level measures of social determinants of health were not available. Neighborhood-level income was available but was not used for matching to limit further loss of sample size, and because area-level median income may not capture the ways in which individual-level income, or other individual-level social determinants, might influence sexual networks.^19^ Furthermore, comparison across groups and the third sensitivity analysis suggest there was no residual confounding by neighborhood-level income. Third, although we included men with a history of bacterial STI, our study eligibility population could be missing men who are at risk of mpox infection but have negligible healthcare access and/or engagement (thus leading to a selection bias). Fourth, data from other studies demonstrate added protective benefit of two doses,^8,10,11^ but we could not evaluate the two-dose regimen because of low second dose coverage during the study period, nor could we evaluate duration of protection. Finally, vaccination could also reduce symptoms and signs of mpox and thus less testing,^20^ which means a higher chance of under-ascertainment of subclinical cases among the vaccinated group, which would lead to overestimation of vaccine effectiveness against infection.

In summary, vaccination with a single dose of MVA-BN vaccine was moderately effective against laboratory-confirmed mpox infection in this population-based study of an evolving outbreak and using a target trial emulation to minimize biases. One implication of our finding is that single-dose vaccination may have been a contributing factor in helping slow down transmission in Ontario in 2022. Achieving high coverage with a full course could be important to prevent resurgence. In the absence of randomized clinical trials, our findings strengthen the evidence that MVA-BN is effective at preventing mpox infection and should be made available to communities at risk.

## Data availability

The dataset from this study is held securely in coded form at ICES. While legal data sharing agreements between ICES and data providers (e.g., healthcare organizations and government) prohibit ICES from making the dataset publicly available, access may be granted to those who meet pre-specified criteria for confidential access, available at https://www.ices.on.ca/DAS.

## Code availability

The full dataset creation plan and underlying analytic code are available from the authors upon request, understanding that the computer programs may rely upon coding templates or macros that are unique to ICES and are therefore either inaccessible or may require modification.

**Correspondence and requests for materials** should be addressed to JCK or SM.

## Acknowledgments

We would like to acknowledge colleagues at Public Health Ontario for access to vaccination data from the Digital Health Information Repository, case-level data from the integrated Public Health Information System, and laboratory data from LabWare. We also thank the staff of Ontario’s public health units who are responsible for mpox case and contact management and data collection. We acknowledge the work of over 30 community-based organizations across Ontario who led vaccine mobilization and implementation with public health units and health care providers.

We thank community advisory board members established as part of the Canada-Africa Mpox Partnership.

This document used data adapted from the Statistics Canada Postal Code^OM^ Conversion File, which is based on data licensed from Canada Post Corporation, and/or data adapted from the Ontario Ministry of Health Postal Code Conversion File, which contains data copied under license from ^©^Canada Post Corporation and Statistics Canada. Parts of this material are based on data and/or information compiled and provided by: Ontario Ministry of Health, Canadian Institute for Health Information, Statistics Canada, and IQVIA Solutions Canada Inc. The analyses, conclusions, opinions and statements expressed herein are solely those of the authors and do not reflect those of the funding or data sources; no endorsement is intended or should be inferred. Adapted from Statistics Canada, Canadian Census 2016. This does not constitute an endorsement by Statistics Canada of this product. We thank IQVIA Solutions Canada Inc. for use of their Drug Information File.

Finally, we thank Nicholas Brousseau and Sara Carazo Perez, Institut national de santé publique du Québec, for helpful discussions regarding methodologies and confounders.

## Patient Consent Statement

ICES is a prescribed entity under Ontario’s Personal Health Information Protection Act (PHIPA). Section 45 of PHIPA authorizes ICES to collect personal health information, without consent, for the purpose of analysis or compiling statistical information with respect to the management of, evaluation or monitoring of, the allocation of resources to or planning for all or part of the health system. Projects that use data collected by ICES under section 45 of PHIPA, and use no other data, are exempt from REB review. The use of the data in this project is authorized under section 45 and approved by ICES’ Privacy and Legal Office.

## Author Contributions

JCK and SM designed the study and analysis plan.

SB, CN, JCK, and LF acquired the data for the work.

CL designed the data extraction and assembly workflow.

CL analyzed the data.

All authors made substantial contributions to the analysis plan and the interpretation of the data. JCK and SM vouch for the data analysis.

CN, SM, and JCK wrote the first draft of the manuscript. All authors critically reviewed the manuscript and decided to proceed with publication.

