## Supplementary Materials for "Effectiveness of one dose of MVA-BN vaccine against mpox infection in males in Ontario, Canada: A target trial emulation"

### **Figure S1: Directed acyclic graph**

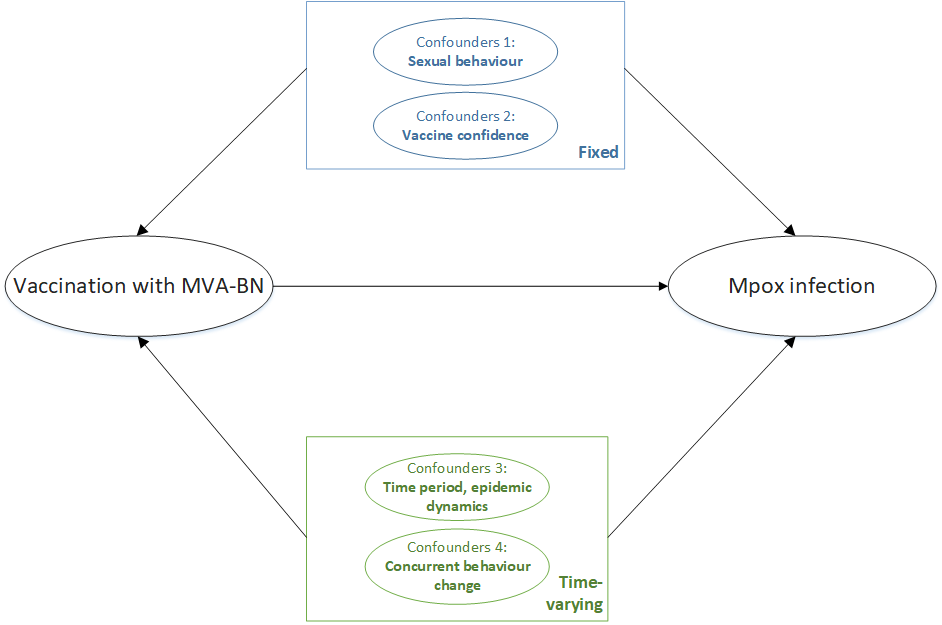

Legend: Directed acyclic graph illustrating fixed and time-varying confounders for vaccination with MVA-BN and risk of mpox infection**;** MVA-BN denotes modified vaccinia Ankara vaccine – Bavarian-Nordic. Fixed confounders were conceptualized as risk of exposure (sexual behaviour) and vaccine confidence, and operationalized by matching on variables. Time-varying, residual confounding related to concurrent behaviour change was examined using the negative tracer outcome (diagnosis of a bacterial sexually transmitted infection, against which MVA-BN should provide no benefit). Time period (or epidemic dynamics) was inherently captured in emulation of the target trial because each vaccinated individual was being compared with a matched unvaccinated individual at the same stage of the evolving outbreak.

### **Supplementary Methods**

#### **Linked data repositories**

Ontario has a single-payer health system that provides universal access to physician and hospital services and laboratory testing for all eligible Ontario residents. For this study, we used population-based databases that capture relevant information within this single-payer system.

MVA-BN was administered as pre-exposure prophylaxis to eligible high-risk groups by local public health units at mass immunization clinics in coordination with community-based organizations, with a smaller number administered via physician clinics. The mass immunization clinics were implemented in drop-in settings, including community-based organizations, with a low-barrier approach to accelerate uptake and maximize access. The low-barrier approach meant that confirmation of eligibility criteria was not required, and any individual who came to the mpox-specific mass immunization clinic could receive the vaccine.

The provincial Digital Health Immunization Repository (DHIR) contains individual-level vaccination data (e.g., product, dose, site, date) entered at the time of administration. Data on post-exposure prophylaxis doses administered by primary care providers were submitted to local public health units for entry into DHIR retrospectively.

All mpox specimens collected by health care providers were submitted to Public Health Ontario laboratories for testing. Laboratory results and demographic and clinical data from the test requisition were obtained from LabWare, Public Health Ontario’s laboratory information system.

Data on syphilis testing were obtained from LabWare and the Ontario Laboratories Information System (OLIS), a provincial repository of laboratory testing results from hospitals laboratories, community-based commercial laboratories, and public health laboratories. Data on prior diagnoses of bacterial sexually transmitted infections (gonorrhea, chlamydia, and syphilis) were obtained from the integrated Public Health Information System (iPHIS), an information system for the reporting and surveillance of Diseases of Public Health Significance.

History of HIV diagnosis was ascertained from the ICES-derived HIV Cohort, which uses an validated algorithm based on physician office visits with HIV as the diagnostic code to identify individuals living with HIV.^1^ Additionally, HIV pre-exposure prophylaxis prescription data were obtained from the Ontario Drug Benefit (ODB) database, which contains information on all publicly-funded prescriptions for: adults aged ≥65 years; recipients of professional home services and social assistance; children and young adults aged ≤24 years and not covered by a private insurance plan; and recipients of the Trillium Drug program, which helps Ontarians who have high prescription drug costs relative to household income.^2,3^

Information on COVID-19 vaccination was obtained from Ontario’s centralized province-wide COVID-19 vaccine registry, COVaxON. Data on influenza vaccines and other publicly funded vaccines received in physician offices were extracted from the Ontario Health Insurance Plan (OHIP) physician billing claims database, while information on influenza vaccines received in pharmacies was extracted from the ODB database.

We obtained age, sex, postal code, and neighborhood-level income quintile from the Ontario Registered Persons Database, a population registry with demographic information for all Ontarians with provincial health insurance. We determined the geographic region (public health unit, which we then collapsed into a smaller number of regions) using the postal code and Statistics Canada Postal Code Conversion File plus (version 7B).

De-identified data extracted from repositories held at Public Health Ontario (Panorama, Labware, iPHIS) were securely shared with ICES, a not-for-profit research institution, and linked to health administrative databases held at ICES using unique encoded identifiers. Linked data were analyzed at ICES.

#### **Matching algorithm**

For each day, starting from June 12, 2022 (beginning of the vaccination campaign) until October 27, 2022 (to ensure each person could have a 30-day observation period), all newly vaccinated persons (15 days after receipt of a first dose) who met the study’s inclusion and exclusion criteria were candidates for matching, including those who had been previously included as unvaccinated controls. These vaccinated individuals were matched in a 1:1 ratio with unexposed (unvaccinated) controls (who fulfilled the inclusion and exclusion criteria and were not previously recruited), in the following ways:

- Age – within five years;
- Geographic region, based on public health unit region (Toronto, Ottawa, Peel/York/Durham/Halton, Hamilton/Niagara/London/Windsor, rest of Ontario) – exact matching;
- Prior HIV diagnosis (yes, no) – exact matching;
- Number of bacterial sexually transmitted infections diagnosed in the prior three years (0, 1, 2, 3, 4 or more diagnoses of any of gonorrhea, chlamydia, or syphilis) – exact matching;
- Receipt of any non-MVA-BN vaccine in the prior year (yes, no) – exact matching.

End of follow-up occurred at the earliest of the following events:

- Occurrence of an outcome event (i.e., testing positive for mpox by PCR)
- Death
- Vaccination of the matched control (for vaccinated individuals)
- Receipt of a second dose of MVA-BN
- Loss of OHIP eligibility (i.e., moved out of province)
- End of the study period (November 26, 2022)

If an unvaccinated individual became vaccinated, the matched pair was censored at the time of the unexposed individual’s vaccination date. The unvaccinated individual who was newly vaccinated was now eligible for inclusion in the study, even if previously selected as a control. This individual was re-matched to an unmatched unvaccinated person and followed until the end of follow-up.

### **Table S1.** Definitions of variables used.

| **Variable** | **Definition** |
| --- | --- |
| Laboratory-confirmed mpox infection | First positive or intermediate test result on real time polymerase-chain-reaction (PCR) testing for orthopox/mpox on samples collected from any body site. |
| Age-group | Age as of June 12, 2023 was determined from the Registered Persons Database, and categorized into the following groups: 18-24 years, 25-29 years, 30-39 years, 40-49 years, and ≥50 years. |
| Geographic region (public health unit region) | Ascertained from Public Health Unit (PHU) information using postal code of residence as recorded in the Registered Persons Database and Statistics Canada Postal Code Conversion File Plus (version 7B). Regions were defined as follows:  Toronto: PHU 95 (City of Toronto Health Unit)  Peel/York/Durham/Halton: PHU 53 (Peel Regional Health Unit), 70 (York Regional Health Unit), 30 (Durham Regional Health Unit), 36 (Halton Regional Health Unit)  Hamilton/Niagara/London/Windsor: 37 (City of Hamilton Health Unit), 46 (Niagara Regional Area Health Unit), 44 (Middlesex-London Health Unit), 68 (Windsor-Essex County Health Unit)  Ottawa: PHU 51 (City of Ottawa Health Unit)  Rest of Ontario: PHU 35 (Haliburton, Kawartha, Pine Ridge District Health Unit), 55 (Peterborough County—City Health Unit), 60 (Simcoe Muskoka District Health Unit), 27 (Brant County Health Unit), 34 (Haldimand-Norfolk Health Unit), 36 (Halton Regional Health Unit), 65 (Waterloo Health Unit), 66 (Wellington-Dufferin-Guelph Health Unit), 38 (Hastings and Prince Edward Counties Health Unit), 41 (Kingston, Frontenac and Lennox and Addington Health Unit), 43 (Leeds, Grenville and Lanark District Health Unit), 57 (Renfrew County and District Health Unit), 58 (The Eastern Ontario Health Unit), 26 (The District of Algoma Health Unit), 47 (North Bay Parry Sound District Health Unit), 49 (Northwestern Health Unit), 56 (Porcupine Health Unit), 61 (Sudbury and District Health Unit), 62 (Thunder Bay District Health Unit), 63 (Timiskaming Health Unit), 31 (Elgin-St. Thomas), 33 (Grey Bruce Health Unit), 39 (Huron County Health Unit), 40 (Chatham-Kent Health Unit), 42 (Lambton Health Unit), 54 (Perth District Health Unit), 75 (Southwestern Health Unit) |
| Number of syphilis tests in past three years | One or more syphilis serological screening tests positive in OLIS or LabWare during the three years prior to or on June 12, 2022 and up to October 27, 2022 categorized as 0, 1, 2, 3, and ≥4. |
| Number of bacterial STIs in past three years | One or more bacterial STI (gonorrhea, chlamydia, or syphilis) diagnosis from iPHIS during the three years prior to or on June 12, 2022 and up to October 27, 2022 categorized as 0, 1, 2, 3, and ≥4. |
| HIV status | An ICES-specific HIV database was used to identify patients with HIV, based on 3 physician claims in 3 years with OHIP diagnostic codes: 042, 043, or 044.^1^ |
| HIV pre-exposure prophylaxis | HIV pre-exposure prophylaxis defined as a dispensation of tenofovir/emtricitabine recorded in the Ontario Drug Benefit (ODB) database, and excluding those prescribed other antiretrovirals within three months of the tenofovir/emtricitabine prescription.^4^ |
| History of receipt of non-MVA-BN vaccines in past one year | COVID-19 vaccines: The date of vaccine receipt from COVaXON.  Influenza vaccines:  Received in physician offices - OHIP feecodes: G590, G591, G592, Q130, Q590, Q690, Q691  Received in pharmacy - ODB billing with any of the following Drug Identification Numbers (DINs): 02420643, 02420783, 02432730, 02473283, 02445646, 02494248, 09857645, 09857646  Other vaccines received in physician offices:  OHIP feecodes: G840, G841, G842, G843, G844, G845, G846, G847, G848, G538, G539 |
| Neighborhood-level income | Household income quintile calculated at the disseminated area (DA) level was used for the neighborhood-level income.  A dissemination area (DA) is the smallest standard geographic area for which all census data are disseminated. A DA generally comprises approximately 400-700 people, but in densely populated cities may contain several thousand people. DAs cover all the territory of Canada.^3^ We assigned subjects to a DA using postal code, as recorded in the Registered Persons Database.  Calculated at the DA level using 2016 Census data by multiplying the median income (before-tax) by the number of households and dividing by the sum of single-person equivalent to obtain income per single person equivalent.^4^ For DAs where median income was unavailable, neighboring DAs were used to estimate income per single person equivalent. DA-based income quintiles were constructed separately for each census metropolitan area or census agglomeration (one or more adjacent municipalities integrated via commuting flows). DAs within each such area were ranked from the lowest average income per single-person equivalent to the highest, and DAs were assigned to five groups, such that each group contained approximately one-fifth the total in-scope population of each area. |

### **Table S2.** Socio-demographic and exposure risk characteristics of the eligible and enrolled study population

|  | **Eligible,**  **n = 3,268 (%)** | **Matched,**  **n = 3,204 (%)** | **SD** |
| --- | --- | --- | --- |
| Inclusion criteria fulfilled |  |  |  |
| ≥ 1 syphilis test and ≥1 bacterial STI, past 1 year | 1,862 (57.0%) | 1,810 (56.5%) | 0.01 |
| HIV PrEP prescription, past 1 year | 1,089 (33.3%) | 1,082 (33.8%) | 0.01 |
| Meeting both criteria | 317 (9.7%) | 312 (9.7%) | 0.001 |
| Age, median (interquartile range) | 35 (29-47) | 35 (29-46) | 0.01 |
| Age group (years) |  |  |  |
| 18-24 | 421 (12.9%) | 418 (13.0%) | 0.01 |
| 25-29 | 451 (13.8%) | 448 (14.0%) | 0.01 |
| 30-39 | 1,146 (35.1%) | 1,122 (35.0%) | 0.001 |
| 40-49 | 573 (17.5%) | 562 (17.5%) | 0 |
| ≥ 50 | 677 (20.7%) | 654 (20.4%) | 0.01 |
| Geographic region |  |  |  |
| Toronto | 2,135 (65.3%) | 2,117 (66.1%) | 0.02 |
| Peel, York, Durham, Halton | 354 (10.8%) | 345 (10.8%) | 0.002 |
| Hamilton, Niagara, London, Windsor | 216 (6.6%) | 212 (6.6%) | 0 |
| Ottawa | 272 (8.3%) | 247 (7.7%) | 0.02 |
| Rest of Ontario | 291 (8.9%) | 283 (8.8%) | 0.003 |
| Neighborhood income quintile |  |  |  |
| 1 (lowest) | 815 (24.9%) | 794 (24.8%) | 0.004 |
| 2 | 758 (23.2%) | 742 (23.2%) | 0.001 |
| 3 | 612 (18.7%) | 604 (18.9%) | 0.003 |
| 4 | 519 (15.9%) | 508 (15.9%) | 0.001 |
| 5 (highest) | 550 (16.8%) | 543 (16.9%) | 0.003 |
| History of HIV diagnosis | 733 (22.4%) | 697 (21.8%) | 0.02 |
| Number of syphilis tests, past 3 years |  |  |  |
| 0 | 133 (4.1%) | 132 (4.1%) | 0.003 |
| 1 | 211 (6.5%) | 208 (6.5%) | 0.001 |
| 2 | 250 (7.6%) | 243 (7.6%) | 0.002 |
| 3 | 288 (8.8%) | 281 (8.8%) | 0.001 |
| ≥ 4 | 2,386 (73.0%) | 2,340 (73.0%) | 0.001 |
| Number of bacterial STIs, past 3 years |  |  |  |
| 0 | 1,166 (35.7%) | 1,158 (36.1%) | 0.01 |
| 1 | 1,003 (30.7%) | 998 (31.1%) | 0.01 |
| 2 | 531 (16.2%) | 522 (16.3%) | 0.001 |
| 3 | 256 (7.8%) | 248 (7.7%) | 0.003 |
| ≥ 4 | 312 (9.5%) | 278 (8.7%) | 0.03 |
| Received any non-MVA-BN vaccines, past 1 year | 3,202 (98.0%) | 3,146 (98.2%) | 0.02 |

SD=standardized difference; STI=sexually transmitted infection (syphilis, chlamydia, gonorrhea); PrEP=Pre-exposure prophylaxis; MVA-BN=modified vaccinia Ankara-Bavarian Nordic vaccine
